# Analyzing the efficacy of a decade-long endeavor: extracurricular medical research training amidst the turmoil of Syria

**DOI:** 10.1101/2024.03.17.24304430

**Authors:** Ibrahem Hanafi, Marah Alsalkini, Kheder Kheder, Maarouf Gorra Al Nafouri, Ahmad Rami Rahmeh, Rami Sabouni

## Abstract

**Objectives:** Medical research in Syria faced significant limitations due to inadequate human and financial resources, exacerbated by the ongoing war. Until recently, the curriculum did not incorporate sufficient training on research skills. Consequently, extracurricular workshops were initiated or utilized nationwide to acquire research-related expertise, aiming to bolster research output. This study aims to characterize and evaluate these training endeavors concerning research-related knowledge, attitudes, barriers, as well as research productivity.

**Methods:** This case-control study encompassed all training initiatives in Syria from 2011 to 2020, both on-site and online. Participants consisted of early career healthcare professionals affiliated with all Syrian universities and were recruited into four equal groups based on the quantity of research projects they undertook and published. Our participants reported and assessed individual extracurricular workshops regarding their gained knowledge, attitudes, and practical skills. These initiatives were compared to curricular training and practical peer-led support regarding these outcomes.

**Results:** The study included 53 on-site and 30 online workshops, displaying diverse distributions and features. Attendance of extracurricular workshops correlated with participants’ knowledge, attitudes, and research productivity (p<0.001). The most effective interventions were massive open online courses and workshops lasting over 12 hours. Extracurricular workshops and peer-led support has comparable effectiveness and associated with higher knowledge, attitudes, and practical skills of their beneficiaries in comparison to curricular training (adjusted p<0.05). Lastly, peer trainers in these workshops exhibited more publications and higher levels of knowledge and attitude than the rest of the sample (adjusted p<0.05).

**Conclusions:** Overall, extracurricular interventions and peer support demonstrated their superiority over curricular training. Despite the varied nature of these workshops and the absence of institutional organization, these approaches exhibited significant potential in enhancing research-related knowledge, promoting positive attitudes, and augmenting research productivity in resource-constrained settings such as Syria.

## INTRODUCTION

Research in Syria faced many challenges before the war, including limited research capacities and financial support, which led to poor quantity and quality of research output (1,2). The situation was exaggerated during the Syrian armed conflict, which severely devastated the medical and educational infrastructure and forced the majority of the medical personnel to flee the country (3,4). Consequently, the remaining physicians had to deal with a flood of cases related and unrelated to war (5). leaving them too little motivation and time to improve their academic skills and conduct scientific research (6). In addition, the war also impacted the psychological wellbeing of healthcare professionals and workplace violence exacerbated the incidence of burn-out syndrome among them, which put them under constant pressure and depleted their mental health, competence, productivity, and work satisfaction (7,8). Moreover, the lack of research opportunities, as well as the poor training and mentoring became more prominent as the country was left with a scarcity of expertise in this field (9,10). It is worth mentioning that evidence-based medicine (EBM) was not formally part of the curriculum in Syrian medical faculties until 2016 (9,11). Hence, the pressing and yet unaddressed requirement remained the implementation of accessible, cost-effective, and sustainable training methodologies aimed at augmenting research-related competencies among Syrian medical professionals (12).

On the other hand, significant efforts started to target this paucity at the different Syrian medical schools and they were mainly led by small teams of residents and medical students (9). Local peer-led workshops arose as an alternative cheap method to assist medical students and residents in this field (13). Initially, these teams leveraged online international resources to enhance their research-related knowledge and skills. This initiative empowered them to autonomously strategize, execute, and publish research studies. Subsequently, they effectively orchestrated workshops across Syria, imparting fundamental principles of medical research to their peers, thereby sharing the expertise and experiences acquired. On the institutional level, these efforts were aided by associations gathering Syrian medical professionals practicing abroad such as the Syrian American Medical Society, who also coordinated advanced online courses regarding EBM and academic writing (14,15). Recently, more basic online workshops were delivered by peers performing research careers abroad (13,16). These online workshops were shown to be as effective as face-to-face workshops on participants’ knowledge (14), but offered valuable advantages like affordability, flexible accessibility (13), and safety especially in war zones,(15) non-governmental controlled areas (17), and during pandemics (18,19), which made them more favorable in countries with limited resources like Syria. The progress made by Syrian efforts on this front paved the way for Syrian doctors to seek further training from international bodies when provided as massive open online courses (MOOCs) (9,16). For instance, remote-sites were liaised for the comprehensive course “the Introduction to the Principles and Practice of Clinical Research (IPPCR)” provided by the American national institutes of health (NIH) in several Syrian medical faculties and in several years. These steps made it possible to leverage from more formal sources of medical research training even for participants in massively destroyed cities like Aleppo (20).

Despite the great efforts spent to achieve all these extracurricular interventions and, when assessed, their proven effectiveness, it is difficult to tell whether they collectively achieved the goals they were aimed at nation-wide. These efforts were not tailored based on comprehensive assessments of the needs and barriers at each phase, location, and field (16). Additionally, they were mostly run by individual or team efforts and lacked cohesive organization and coordination, which hindered both the learning and teaching processes in the long run and challenged any comparison of the efficacy of the different efforts. Consequently, it is still to be proved that the Syrian medical research synthesis significantly improved in quality and quantity due to these initiatives, as most of the research articles published from Syria are still mostly case reports and cross-sectional studies and when they do not end up unpublished, they mostly get published in low impact journals. Therefore, this study aimed to characterize all extracurricular training opportunities utilized by early career Syrian healthcare workers during the first decade of the Syrian war and to assess their impact on attendees’ knowledge, attitudes, perceived barriers, as well as their research productivity.

## METHODS

### The identification of extracurricular events targeting medical research

We identified all interventions targeting research capacity building for Syrian healthcare professionals during the period 2011-2020. This was performed through interviews with trainers and trainees from different academic levels and at all Syrian medical schools. We additionally performed a comprehensive search for event invitations on social media platforms, particularly in groups and channels that target Syrian healthcare professionals and medical and paramedical students around the country. The final list of workshops included all extracurricular in-house training that took place at any of the Syrian medical schools as well as those offered online by international associations or platforms like the American National Institute of Health (NIH) and Coursera. We then extended our interviews to retrospectively define the features of each workshop including its city, year, trainers (professionals or peers), length (approximate number of training hours), size (approximate number of attendees), and focus (research methodology, critical appraisal, academic writing, statistical analysis, systematic reviews, and case reports). Online workshops were also classified into international workshops (i.e., offered by the NIH, Cochrane, Coursera, and Stanford school of Medicine) and Syrian workshops (i.e., offered by the Syrian American Medical Society and Syrian peers abroad). All of the international workshops were delivered in English, while most of the Syrian workshops were presented in Arabic. During data collection, participants also had the possibility to add missing workshops and provide details about their features (e.g., year, city, and duration).

### Study design and population

This study had a case-control design, and its sample population is identical to the one reported in our previous article (21). The recruitment for data collection took place between 05.01.2021 and 06.07.2021. Briefly, we communicated the questionnaire individually to all undergraduate students, postgraduate students, residents, and fresh specialists in all Syrian medical and paramedical faculties, who have at least one published article in a peer-reviewed journal. These participants were grouped into two equal arms, the first with one publication and the second with two or more publications. We then recruited two equal control groups, one with participants who contributed to at least one research study (without any publication) and the second with participants who had no previous research contributions. We maintained equal arms at every university and academic level (under- and postgraduates). Syrian specialists and postgraduate students who are currently practicing outside Syria but received their medical training in one of the Syrian universities were not excluded. Participants in their first or second year of undergraduate studies or after more than one year from the end of their specialization were excluded. Further details regarding the ethical approval, data collection, and sample characteristics can be found in our previous article (21); which also reported participants’ self-reported research-related knowledge, attitudes, and perceived barriers. Briefly, the ethical approval for this study (30/2020) was obtained from the institutional review board of Damascus University on 19.01.2021. All participants provided their written informed consent for the publication of their anonymous data before their participation.

### The data collection tool

The questionnaire developed first asked the participants to identify all the workshops and trainings they took part in by selecting them from a list that contained the name, year, and city of each workshop. In the next step, participants had the possibility to refer to their first, second, and third best workshops in terms of knowledge and skills gained from it. For each of these one to three workshops, participants also assessed the gained knowledge, attitudes, and practical practice on a scale from one to ten, where one stood for poor benefit and ten stood for outstanding benefit. In order to compare these evaluations to other methods of gaining research-related knowledge and skills, we asked the participants to perform the same evaluation for undergraduate and postgraduate curricular research training as well as the practical support they received from peers they worked with on specific research projects. As most of the in-house training was peer-led, we supplemented the questionnaire with a section asking the participants to identify the workshops they participated in training. This information was used afterwards to compare the cohort of these peer trainers to the rest of the sample in terms including knowledge, attitudes, and publications.

### Data analysis

The data was exported to Microsoft Excel 365 version 2011 (year 2020) then analyzed using the Statistical Package for the Social Sciences version 23.0 (SPSS Inc., Chicago, IL, United States). Details about the presentation and statistical analysis of the demographic characteristics, knowledge, attitudes, and barriers including association and correlation tests can be found in the first article (21). In addition, we used different logistic regression tests for models that predicted conducting and publishing research studies as well as number of extracurricular workshops attended and research knowledge and attitudes. For these analyses, regression coefficients and their 95% confidence intervals (CI) were reported as measures of association together with their P values.

As most of the participants with multiple publications attended several workshops with various features, we could not link higher research productivity (i.e., number of conducted and published research projects) to specific workshops characteristics. However, to sidestep this obstacle, we grouped participants repeatedly based on the dominant features in the workshops they attended. These groups of participants were then compared to draw conclusions about features responsible more for higher research conduction. For instance, if more than half of the workshops attended by a participant were delivered by peers, he was included in the group of peer-led training and otherwise he would be in the professional-led training group. The same applied when more than half of the training attended by the participant happened online, originated from international organizations, focused on research methodology, took place after 2017, had more than 12 training hours, and recruited more than 40 participants.

To limit the number of workshops presented, we excluded those that were attended by less than four participants from our population. To mitigate the possibility of over- or underestimating the rank or the self-assessed benefit from any workshop, we also excluded all workshops that received less than three evaluations (by three participants). For the rest, we allocated points based on the ranking the participants gave to each workshop, where workshops ranked first, second, and third best got three, two, and one points, respectively. To adjust the rankings for the number of attendees, we divided the sum of the points of each workshop by the number of participants in our sample who reported attending this workshop. On the other hand, the gained knowledge, attitudes, and practical practice for each workshop were averaged among entries.

An Alpha value of 0.05 was used as a cutoff to determine statistical significance after Bonferroni correction for multiple comparisons. Figures were created using Microsoft Excel 365 version 2011 (year 2020) and the Statistical Package for the Social Sciences version 23.0 (SPSS Inc., Chicago, IL, United States), while illustrations were designed using Adobe Illustrator (year 2017) and Microsoft PowerPoint 365 version 2011 (year 2020).

### Patient and Public Involvement

The target audience for this project encompasses medical under- and postgraduates, and this report presents their assessment of the needs and challenges faced towards better research productivity. It is important to note that this project was solely executed by a team comprising these individuals, ensuring that the research directly addresses their needs and interests.

## RESULTS

### The demographics of extracurricular medical research workshops

Fifty-six in-house workshops that targeted research-related skills were identified during the study period. Our workshops’ list additionally included 53 online workshops that were accessible to Syrian healthcare personnel to learn about medical research. Of which, three in-house and 23 online workshops were attended by less than four of our participants and were, therefore, excluded from the sample. The few in-house workshops that took place before 2017 were concentrated in Damascus. Afterwards, workshops increased in number and extended to the other two major Syrian medical schools (i.e., Aleppo, and Latakia). In general, the workshops varied in the length of training, number of trainees, and covered topics. However, it can be noted that the most common trained topic of in-house workshops was research methodology (26; 49%), and these workshops had larger size than the ones focusing on other research skills. It is also worth mentioning that the topics covered were not homogenously distributed across the years (e.g., most systematic review workshops happened between 2016 and 2017). Additionally, the majority of in-house workshops (n=45; 85%) were conducted by peers, leaving only seven professional-led workshops, which focused mainly on research methodology. On the other hand, the majority of online workshops were delivered by professionals as MOOCs (n=22; 71%), and they mostly had more than 12 training hours. However, we could still isolate a group of online workshops delivered by Syrians, which were classified into professional-led trainings conducted by the SAMS and peer-led ones (Fig 1).

**Fig 1:**
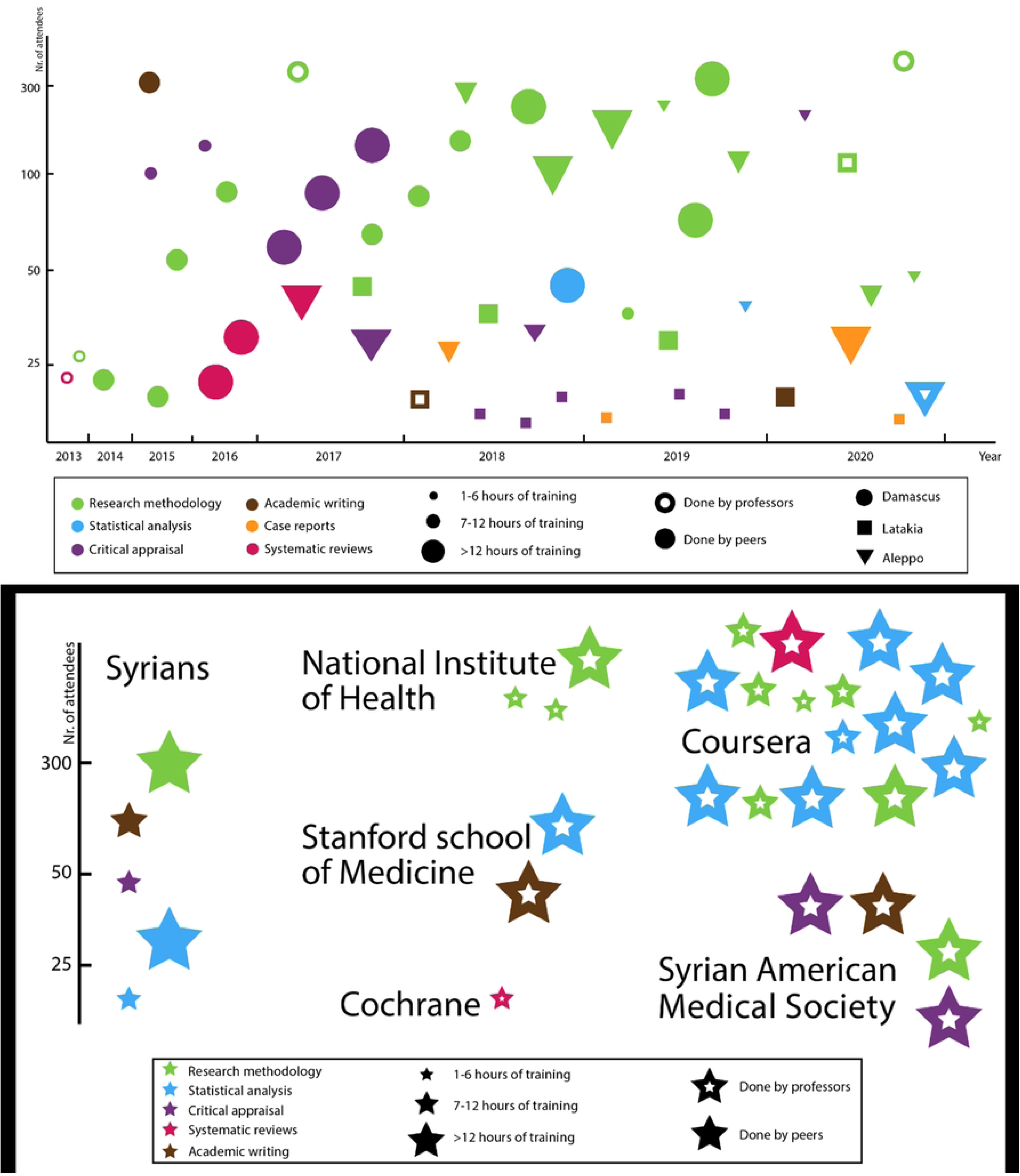
Inhouse and online training sources on medical research. Panel a illustrated inhouse trainings while panel b illustrates online sources of training.

The vast majority of these extracurricular workshops were offered for free and to all under- and postgraduate students as well as specialists. Nevertheless, we found that participants affiliated with the University of Aleppo attended more workshops compared to participants in Damascus and Latakia (*P*<0.001). Furthermore, participants who had better internet connectivity or higher English competence also attended more workshops (S. table 1).

### The effectiveness of extracurricular medical research workshops

After excluding the participants who never attended any research focused extracurricular workshop, we found a significant moderate positive correlation between the number of extracurricular trainings attended and the number of research projects conducted. The number of attended trainings also correlated with the number of publications as well as the total scores of knowledge and attitudes (Fig 2). Moreover, we entered the number of trainings attended together with the total scores of knowledge, attitudes, and barriers, as well as the academic score in a stepwise multiple regression model to predict the number of research projects conducted or published for each participant. This analysis showed that the total knowledge score could independently predict research conduction and publication similarly to our previous article.(21) However, the number of attended extracurricular workshops had an independent added change of 3.6 % to the R2 (P<0.001) specifically in the prediction of number of conducted research studies (S. table 2).

**Fig 2:**
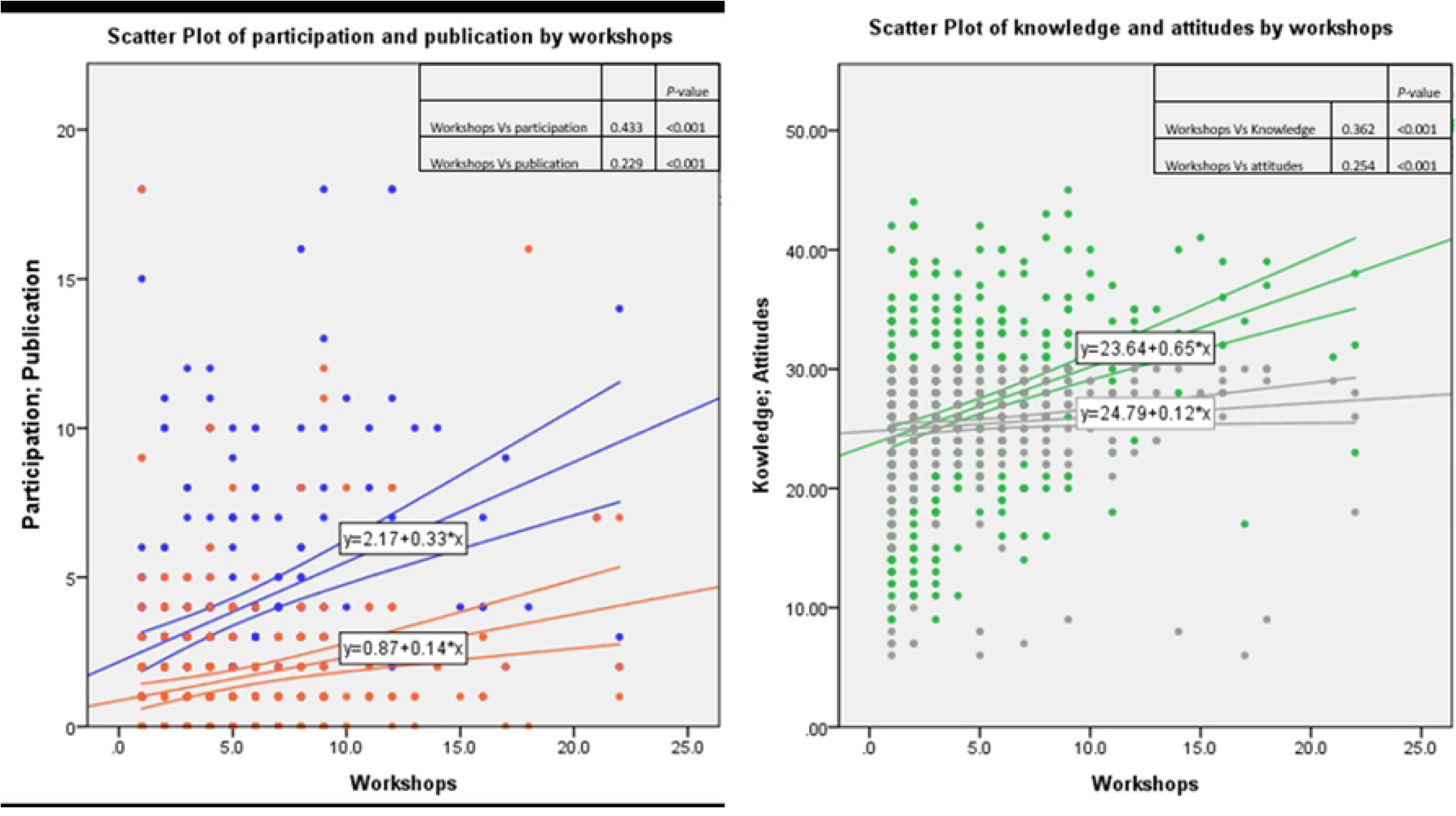
The impact of the number of workshops attended on research productivity as well as the self-reported knowledge and attitudes towards medical research. Panel a represents the correlations of conduction (blue) and publication (orange) of research studies, while panel b represents the same for the total knowledge (green) and attitudes (grey) scores. The correlations were estimated based on Spearman’s rho methods. Participants who never attended any workshop were excluded (n=452 for participation and panel b; n=364 for publication).

### Comparing the features of medical research extracurricular workshops

MOOCs and workshops that had more than 12 training hours were associated with a higher number of conducted and published studies as well as higher knowledge scores than Syrian-led trainings and shorter workshops, respectively (Fig 3 and Fig 4). These findings were confirmed by stronger correlations between the number of workshops attended with these features on the first side and research productivity, knowledge, and attitudes on the other side (S. table 3). However, as these two features significantly coexist (i.e., most MOOCs had more than 12 training hours), we ran an ordinal logistic regression that confirmed the independent contributions of these two features to the higher research productivity (S. table 4). Furthermore, to isolate the added value of these two features, we added the total knowledge score (i.e., the best predictor of publications) to the same logistic regression. Interestingly, MOOCs lost their independence in predicting the number of publications while the length of the workshops retained its independent effect (S. table 5). On the other hand, participants who attended more professional-led trainings also conducted more research projects and had higher knowledge than those who attended more peer-led trainings. This difference was evident on the correlation analysis but did not show independent contributions to the predictions in the regression analyses.

**Fig 3:**
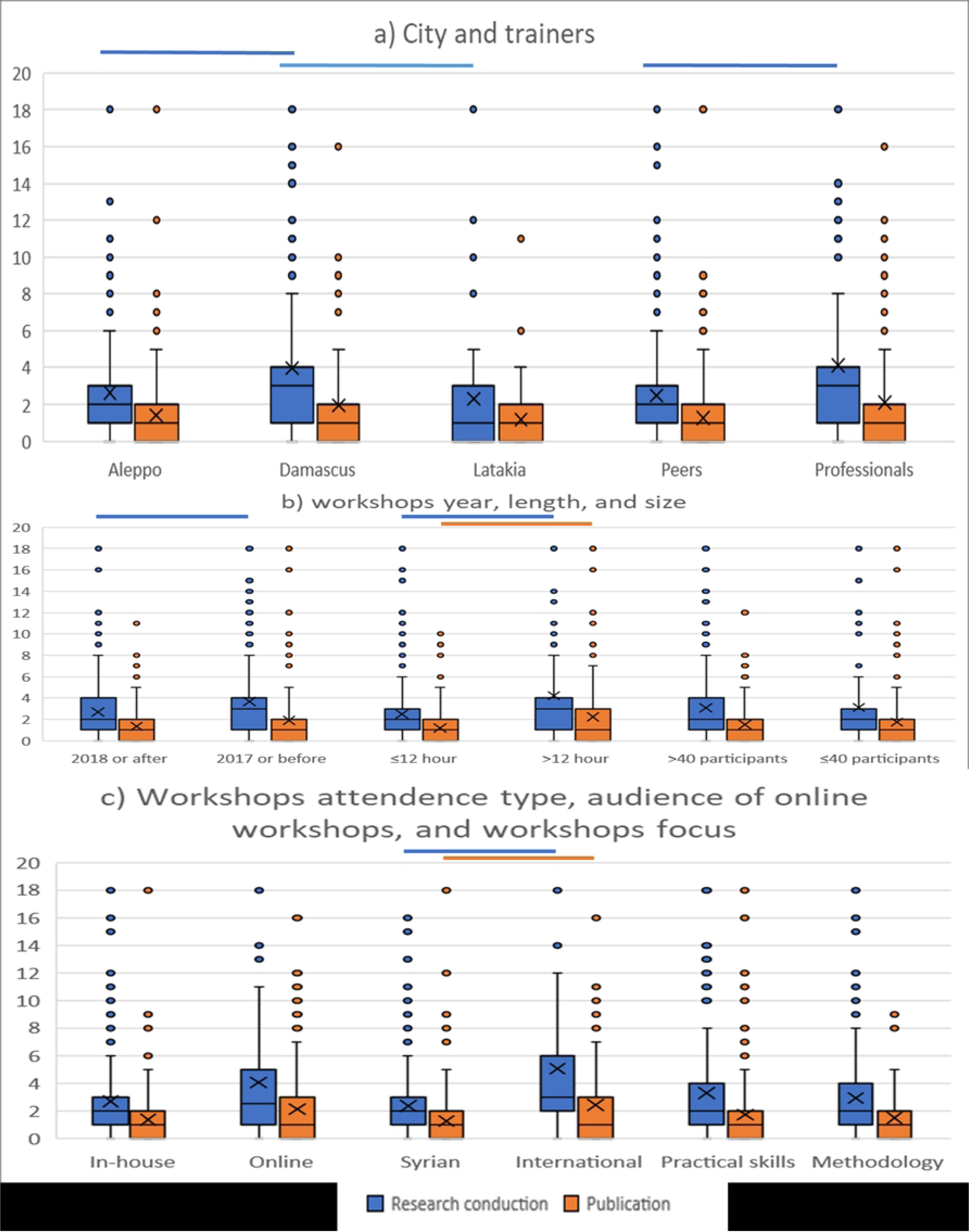
The impact of workshops features on research conduction and publication. The bars refer to statistically significant differences in Mann-Whitney U test after Bonferroni correction for multiple comparisons (P=0.05/8=0.006); the boxes depict interquartile ranges; the horizontal lines in the boxes present the medians; the X signs show the means; the extending vertical lines illustrate the ranges; and the dots that exceed the line represent outliers.

**Fig 4:**
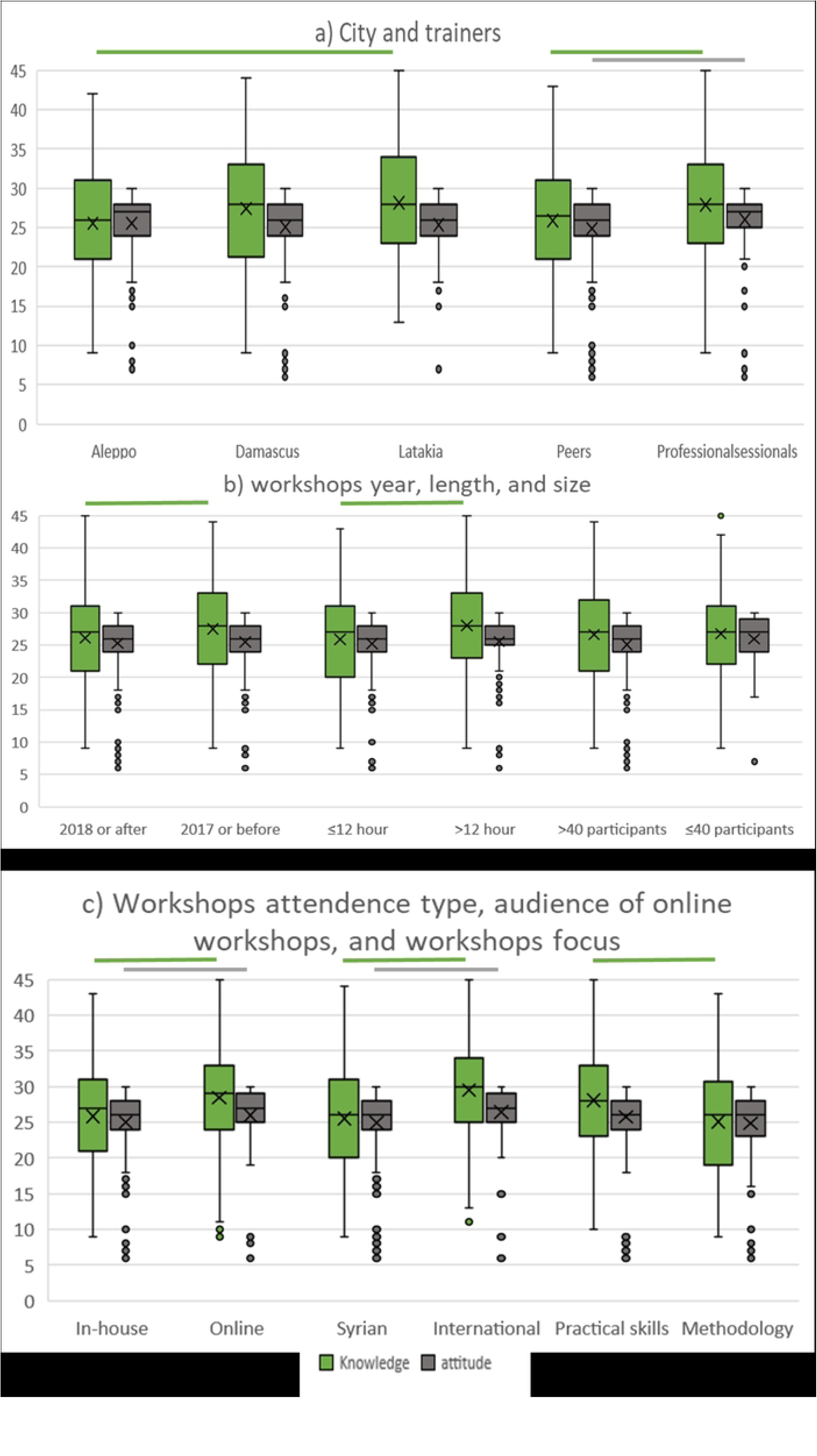
The impact of workshops features on participants’ knowledge, and attitudes towards research. The bars refer to statistically significant differences in Mann-Whitney U test after Bonferroni correction for multiple comparisons (P=0.05/8=0.006); the boxes depict interquartile ranges; the horizontal lines in the boxes present the medians; the X signs show the means; the extending vertical lines illustrate the ranges; and the dots that exceed the line represent outliers.

### Participants’ evaluation of extracurricular medical research workshops, curricular training, and peer support

Three-hundred-fifty-six participants of our sample indicated the best one to three extracurricular trainings they attended then self-evaluated their gained knowledge, attitudes, and practical skills from each one of them. After excluding the workshops that received less than three individual evaluations, only 54 workshops remained for the analysis. These assessments showed that averaged evaluations of the one to three best extracurricular workshops refer to higher gained knowledge, attitudes, and practical practice than the average of under- and postgraduate official curricular training. It also showed that practical peer support was rated as high as the extracurricular workshops and was also significantly higher than curricular training on the three scales (S. Fig 1). We then ran a correlation analysis between the self-evaluated gained knowledge and attitudes from each of the three training sources with the total knowledge and attitudes scores. The correlations confirmed our findings as they were significant for extra-curricular workshops and peer support but not for the curricular training. Additionally, the estimation of the gained knowledge, attitudes, and practical practice from peer support correlated significantly with research productivity, which did not apply to evaluations regarding curricular training. Meanwhile, the same correlations regarding extracurricular workshops did not survive correction for multiple comparisons (S. table 6).

To compare the evaluations of gained knowledge, attitudes, and practical practice among the different features of the workshops, we averaged the assessments of each workshop across participants. Then, we compared the groups of workshops for each gained outcome. However, the only difference that survived the correction for multiple comparisons was the higher practical skills gained from MOOCs when compared to other online workshops (S. Fig 2).

### Training on extracurricular medical research workshops

Seventy-five participants in our sample trained in at least one of the studied peer-led workshops; most of them (n=62, 82.7%) trained in less than three workshops, and 49 of them (65.3%) were affiliated with the University of Aleppo (S. table 7). Participants who trained in workshops reported higher numbers of research projects conducted and published as well as greater knowledge and attitudes compared to those who did not train on any workshop regardless of whether they attended or did not attend any workshop. Trainers also showed higher English competence and attended more workshops compared to those who did not participate in training any workshop (Fig 5). Among these five variables (i.e., research conduction, publication, knowledge, attitudes, and English level), the number of research projects conducted, and knowledge and attitudes scores were isolated as significant independent predictors of the variability between those who trained and did not train on workshops in a binary logistic regression analysis (S. table 8).

**Fig 5:**
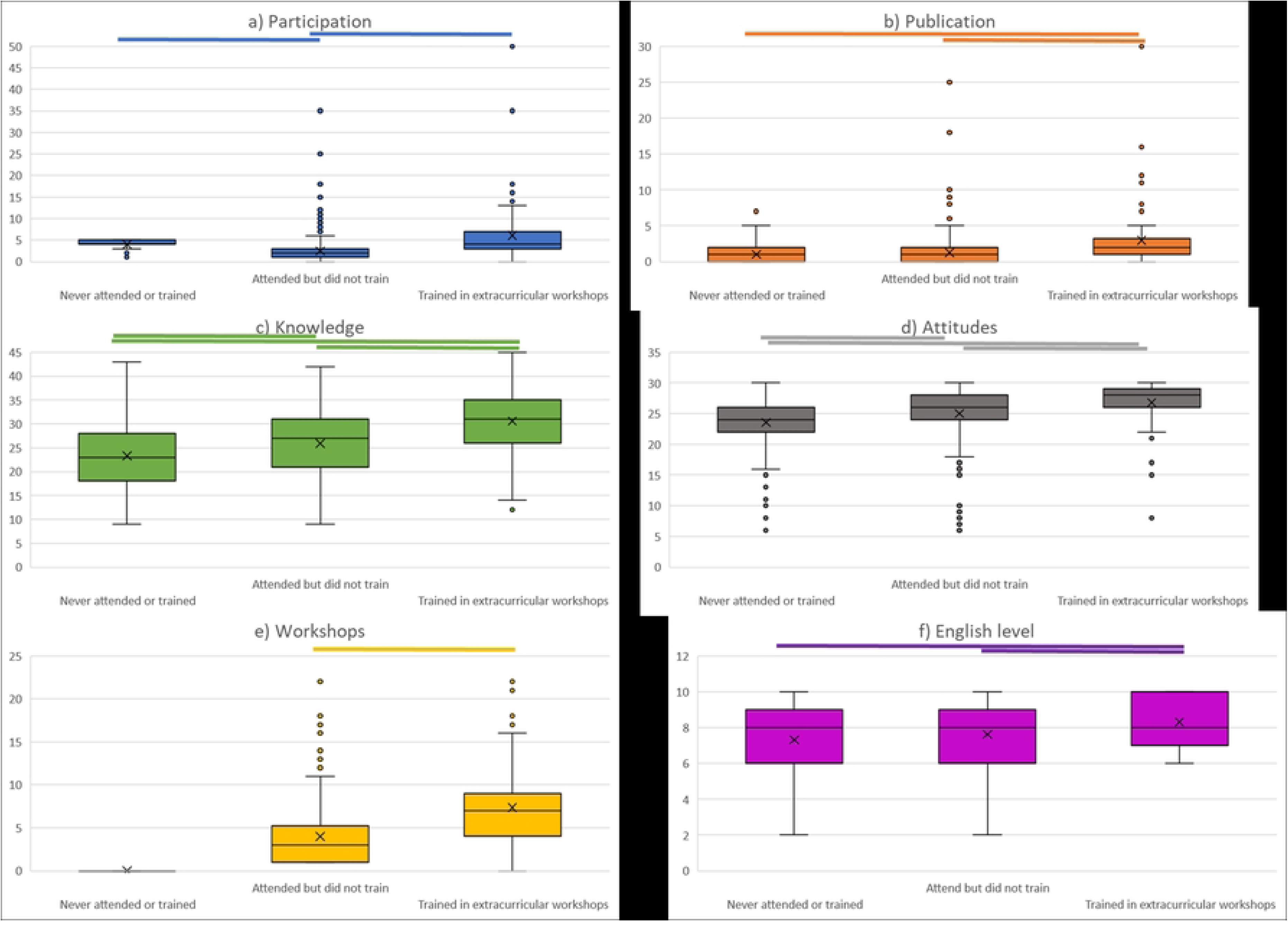
Comparison of the demographic characteristics and research contributions between peer trainers and attendees of research related trainings as well as those who never attended any of these trainings. The bars represent statistically significant differences in the post-hoc Dunn-Bonferroni-Test pairwise comparison (adjusted P<0.05); the boxes depict interquartile ranges; the horizontal lines in the boxes present the medians; the X signs show the means; the extending vertical lines illustrate the ranges; and the dots that exceed the line represent outliers.

## DISCUSSION

Despite the unprecedented humanitarian crisis in Syria, Syrian students sought to develop their research skills by attending numerous in-house and online workshops conducted through Syrian efforts or provided by international organizations. This study was novel in providing a broad overview of the characteristics and effectiveness of all these interventions used during the first ten years of the Syrian armed conflict. We first proved that attending more of these workshops was associated with higher research knowledge, attitudes, and productivity. Then, we showed that self-reported research knowledge had particularly the strongest correlation with the number of attended workshops and was also the strongest predictor of the number of conducted and published research projects. Additionally, we could, using participants’ evaluations, show that these extracurricular interventions were more effective than curricular training and as valuable as practical peer support during ongoing research projects. Finally, we presented a scheme summarizing all the modifiable personal factors that play a role in improving or hindering research productivity in our limited resources settings (Fig 6).

**Fig 6:**
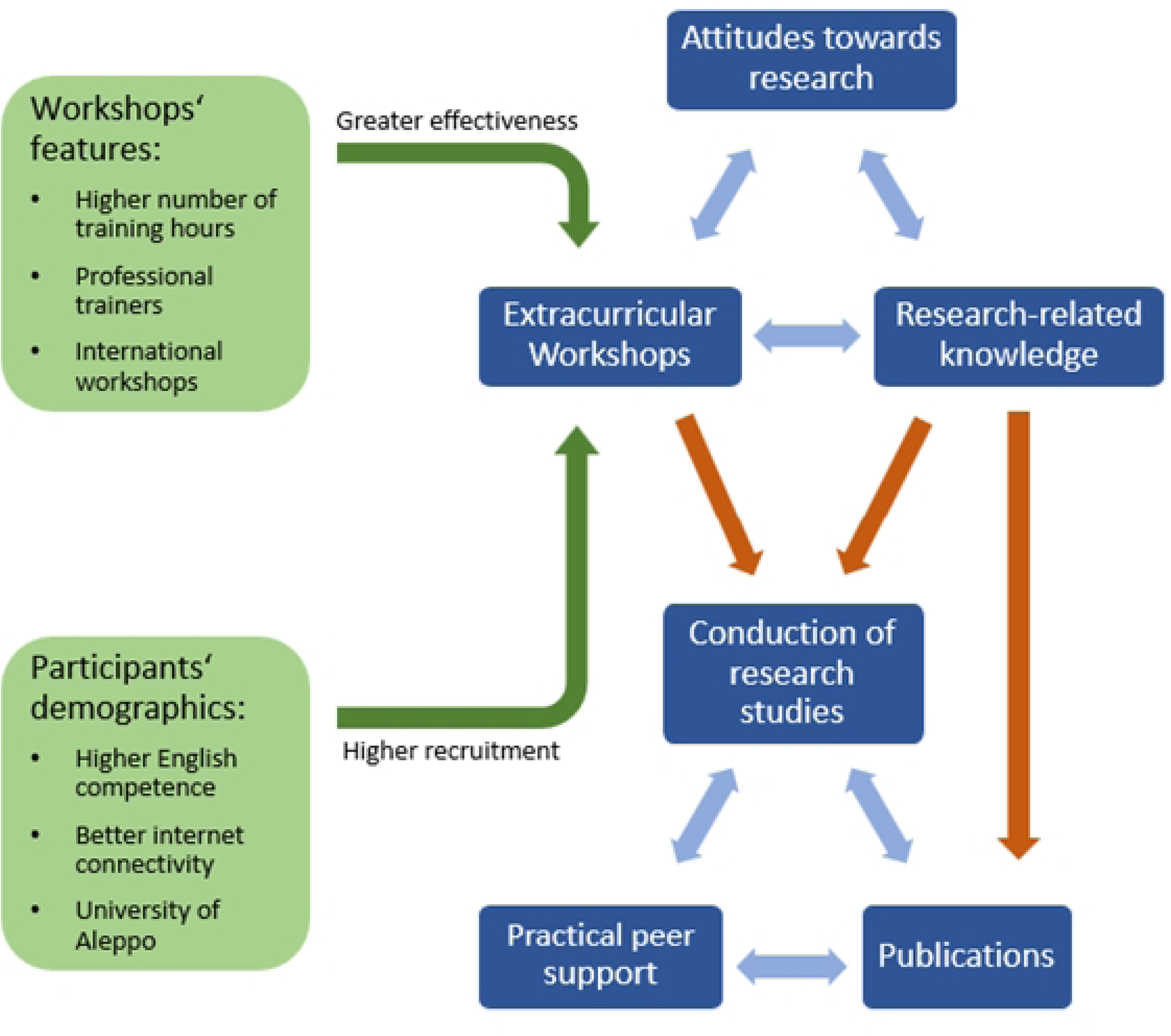
A scheme depicting all the investigated factors that affect research productivity among early career health professionals as well as the interrelations among them. The blue arrows represent significant positive correlations; the orange arrows illustrate significant independent predictors with the dependent variable ahead of the arrow; and the green arrows depict significant association with higher effectiveness or recruitment of the extracurricular trainings.

Even in countries where research training is deeply rooted as a part of the curriculum, research workshops seemed to be the most effective way of training (22). These workshops are also widely recognized as an effective tool for promoting medical research in low- and middle-income countries (LMICs) (23–25). In Syrian, they were suggested as an attainable alternative to address the shortcomings of the research curriculum (10,16). Previous reports assessing individual interventions that targeted research skills including EBM and academic writing in Syria highlighted the knowledge-enhancing effects of their workshops (11,13,14,26), or at least improvement limited to attendees’ confidence (27), or perceived skills (15). Additionally, a recent cross-sectional study on Syrian residents suggested that attending more trainings on medical research correlates with better practices of EBM (28). We similarly proved that attending more research workshops has a significant positive impact on research-related knowledge. Although this finding comes in line with previous reports, it uniquely generalizes their findings employing a novel comprehensive assessment that pooled all eligible interventions in the study period and recruited a large sample size from several universities and academic levels.

Our findings also confirmed that participants who attended more workshops reported higher attitudes towards research, which was described as a major outcome of one workshop targeting EBM.(26) Notably, attitudes towards research were less strongly correlated with workshops attendance compared to knowledge. We can attribute that to the fact that medical personnel in Syria, even those who have never conducted any research project, generally reported high attitudes towards research (10,16), and this does not apply to the poor research-related knowledge repetitively reported (21,28,29). The weak impact of workshops on attitudes can also be due to the limited effectiveness of workshops in addressing certain research barriers, such as language, internet connection, and time availability (10,11,16,29), thus, hindering attitudes strengthening towards medical research.

Despite that, when it comes to studying the improvement in research productivity, trainings on research skills can achieve it in several pathways (Fig 6). Our previous article established that self-reported knowledge explains much of the variability in the number of conducted and published scientific papers (21). The current study also proves that attending research-related workshops as an independent factor has a positive additional value on research conduction. This can be attributed to the development of leadership and time management skills through extracurricular activities (30) and the possibility to engage more effectively in the research community during such trainings (31). The extracurricular workshops in Syria were also suggested to help in establishing research networks that foster research knowledge and participation, in addition to enhancing students’ understanding of research materials (10,20). The higher research productivity we found among participants who attended longer workshops supports this logic, as the time by itself may offer a higher probability to the establishment of the aforementioned research networks. Although, the literature suggests that shorter courses can be of higher feasibility and effectiveness (12,32), they might not offer further mentorship after the training (33), possibly suggesting a lower impact on research productivity, which meets our findings.

Although MOOCs and traditional learning showed similar effectiveness in medical education by means of a meta-analysis (34), MOOCs remain a valuable, and sometimes the only, asset in LMICs to acquire the needed knowledge and skills to conduct research projects (35). In line with that, our study found that participants who attended MOOCs reported higher knowledge and research productivity than those who attended other workshops. This can be explained by the findings of Li et al, which highlighted that MOOCs attendees tend to have higher motivation to learn (36). The open-structure of MOOCs also allows students to design their individual learning objectives depending on their needs (37). Therefore, we can speculate that participants who had running projects preferably attended MOOCs to address the specific knowledge gaps they faced during the progress of their projects, which could be the reason behind the higher knowledge and research conduction we found among MOOCs attendees. A similar pattern was evident in an academic writing workshop, where participants who practiced research skills through writing assignments gained higher objective knowledge than those who did not achieve these assignments (13).

Although the investigated workshops were in general effective in improving research knowledge, attitudes, and productivity, it is difficult to believe that Syrian healthcare personnel leveraged from the full capacities of these interventions. It was quite evident that these interventions lacked a consistent pattern across the different cities and varied in terms of content, delivery, and targeted group(s). This lack of organization can be attributed to the fact that they were in most cases driven by the needs in their proximity, which suggests a significant room for improvement if they become tailored based on up-to-date representative assessments of the needs and barriers towards research (10,13,16,21). On the other hand, we assume that the lack of institutional collaborations constrained the amplitude of benefit, especially from in-house workshops, to specific groups and locations. Therefore, it is important to emphasize the role of the concerned faculties and ministries in organizing research training efforts. This is particularly relevant in developing countries, where personnel with decision-making capacities are less involved in the scientific progress (38).

Nevertheless, this study emphasized the role of peers in pushing the wheel of research training in Syria (9). The vast majority of in-house trainings, as well as some online workshops, in our analysis relied heavily or completely on under- or postgraduate peer trainers with wider research experiences and higher research-related knowledge and attitudes. This role of peers was particularly underlined in countries grappling with significant training gaps and depletion of professionals, as reported in the Syrian case (10,11,13,16). The reliance on peers in such settings is both justified and indispensable, as peer-led training presents a practical and affordable solution that alleviates the burden on professors and faculty members (39,40). Superiority of peer training might also be argued, as one study also reported negative experiences (e.g., uncomfortable interaction) in extracurricular activities delivered by faculty members (30). Notably, the support from peers extended beyond extracurricular trainings to personalized peer-to-peer assistance during ongoing research projects (9,16). Our study further underscored that these two forms of peer-support are of comparable value in the gained knowledge, attitudes, and practice. Interestingly, both of these methods were significantly more valuable to beneficiaries than the previously deemed substandard curricular research training (10,11,16), suggesting them as an established solution for the dearth of expertise in Syria (13).

### Limitations

In spite of the strengths that reside in our sample and methodology, our findings still provide proof of association rather than causality. For instance, it might be argued that participants with higher knowledge, attitudes, and more research projects are those who attend workshops training research skills. However, this remains unlikely for two reasons. First, this explanation does not justify how high our participants evaluated the best workshops they attended (S. Fig 1). Second, a significant group of participants had multiple research projects despite declaring that they never attended any workshop training research skills (Fig 5). On the other hand, one systematic review suggests that increased knowledge may be driven by medical students’ motivation to achieve higher grades on curriculum-based assessments (41). This explanation does not apply to our sample, as several reports described how Syrian researchers lack the institutional and curricular motivation to perform research (10,11,16). Additionally, the academic scores of our participants did not correlate with any measure of attitudes towards research (21). Moreover, the tool we used remains subjective and lacks the objectivity in assessing research-related knowledge in particular. It might have also been affected by recall bias as some of the trainings were ten years old by the time of data collection. Finally, as many participants attended a mixture of workshops of different characteristics, it was quite difficult to purely assess the impact of each of their characteristics individually. However, we believe that the clustering methods we used in addition to the self-evaluation of the best workshops are the best feasible approaches to overcome this obstacle.

### Conclusion

The extracurricular interventions targeting medical research paucity in Syria have proven successful in helping Syrian early-career medical researchers to find their voice (27). Regardless of the dreadful circumstances of war, these workshops increased research attitudes, knowledge, skills, and productivity, and facilitated engagement in the research community. Local peers and international professionals played crucial roles in this progress; however, these efforts still lack the institutionally-organized framework that can be achieved by joining forces with medical faculties.

## DECLARATIONS

### Acknowledgements

The authors are grateful to Luma Haj Kassem and Ahmad Naem for their support in data collection.

### declaration of interest

None of the authors have any conflict of interest that is relevant for this project.

### Contributions

IH conceptualized the project. IH, MA, KK, ARR collected the data. IH, RS, and ARR analyzed and interpreted the data. IH, MA, KK, and MGN prepared the manuscript. IH supervised the project. All authors revised and approved the final version of the manuscript.

### Ethical approval and consent for publication

The ethical approval for this study (30/2020) was obtained from the institutional review board of Damascus University on 19.01.2021. All participants provided their written informed consent for the publication of their anonymous data before their participation.

### Data availability statement

The data supporting this manuscript is available and can be provided upon reasonable request to the corresponding author.

### Funding

None of the authors received any funding from any agency that is related to the work on this project.

## PRACTICE POINTS

Syrian extracurricular training initiatives positively impact knowledge, attitudes, and research productivity among early career healthcare professionals.

Expanding extracurricular initiatives could offer a maintainable method to significantly enhance research capacity in resource-constrained settings.

Massive open online courses and longer-duration trainings are more effective in achieving these outcomes.

Policymakers and educators may incorporate these initiatives to optimize research outcomes despite limited resources.

## Notes

### Competing Interest Statement

The authors have declared no competing interest.

### Funding Statement

The author(s) received no specific funding for this work.

### Author Declarations

The ethical approval for this study (30/2020) was obtained from the institutional review board of Damascus University on 19.01.2021. All participants provided their informed consent for the publication of their anonymous data before their participation.

## REFERENCES

1. Matar HE, Almerie MQ, Adams CE, Essali A. Publications indexed in Medline and Embase originating from the Syrian Arab Republic: a survey. East Mediterr Health J Rev Sante Mediterr Orient Al-Majallah Al-Sihhiyah Li-Sharq Al-Mutawassit. 2009;15(3):648–52.

2. Diab MM, Taftaf RMO, Arabi M. Research productivity in Syria: Quantitative and qualitative analysis of current status. Avicenna J Med. 2011 Jul;1(1):4–7.

3. Sahloul MZ, Monla-Hassan J, Sankari A, Kherallah M, Atassi B, Badr S, et al. War is the Enemy of Health. Pulmonary, Critical Care, and Sleep Medicine in War-Torn Syria. Ann Am Thorac Soc. 2016 Feb;13(2):147–55.

4. Coutts A, Fouad FM. Response to Syria’s health crisis--poor and uncoordinated. Lancet Lond Engl. 2013 Jun 29;381(9885):2242–3.

5. Alahdab F, Omar MH, Alsakka S, Al-Moujahed A, Atassi B. Syrians’ alternative to a health care system: “field hospitals.” Avicenna J Med. 2014 Jul;4(3):51–2.

6. Fouad FM, Sparrow A, Tarakji A, Alameddine M, El-Jardali F, Coutts AP, et al. Health workers and the weaponisation of health care in Syria: a preliminary inquiry for The Lancet-American University of Beirut Commission on Syria. Lancet Lond Engl. 2017 Dec 2;390(10111):2516–26.

7. Mohamad O, AlKhoury N, Abdul-Baki MN, Alsalkini M, Shaaban R. Workplace violence toward resident doctors in public hospitals of Syria: prevalence, psychological impact, and prevention strategies: a cross-sectional study. Hum Resour Health. 2021 Jan 7;19(1):8.

8. Alhaffar BA, Abbas G, Alhaffar AA. The prevalence of burnout syndrome among resident physicians in Syria. J Occup Med Toxicol Lond Engl. 2019;14:31.

9. Al Saadi T, Abbas F, Turk T, Alkhatib M, Hanafi I, Alahdab F. Medical research in war-torn Syria: medical students’ perspective. Lancet Lond Engl. 2018 Jun 23;391(10139):2497–8.

10. Turk T, Al Saadi T, Alkhatib M, Hanafi I, Alahdab F, Firwana B, et al. Attitudes, barriers, and practices toward research and publication among medical students at the University of Damascus, Syria. Avicenna J Med. 2018;8(1):24–33.

11. Alahdab F, Firwana B, Hasan R, Sonbol MB, Fares M, Alnahhas I, et al. Undergraduate medical students’ perceptions, attitudes, and competencies in evidence-based medicine (EBM), and their understanding of EBM reality in Syria. BMC Res Notes. 2012 Aug 12;5:431.

12. Busse CE, Anderson EW, Endale T, Smith YR, Kaniecki M, Shannon C, et al. Strengthening research capacity: a systematic review of manuscript writing and publishing interventions for researchers in low-income and middle-income countries. BMJ Glob Health. 2022 Feb;7(2):e008059.

13. Hanafi I, Kheder K, Sabouni R, Gorra Al Nafouri M, Hanafi B, Alsalkini M, et al. Synchronicity and peer-reviewed assignments in online scientific writing training with limited resources: a four-arm blinded randomized controlled trial with one-year follow-up [Internet]. 2022 [cited 2023 Jun 3]. Available from: https://www.researchsquare.com

14. Sabouni A, Bdaiwi Y, Janoudi SL, Namous LO, Turk T, Alkhatib M, et al. Multiple strategy peer-taught evidence-based medicine course in a poor resource setting. BMC Med Educ. 2017 May 4;17(1):82.

15. Alahdab F, Alabed S, Al-Moujahed A, Al Sallakh MA, Alyousef T, Alsharif U, et al. Evidence-based medicine: a persisting desire under fire. Evid Based Med. 2017 Mar;22(1):9–11.

16. Hanafi I, Haj Kassem L, Hanafi M, Ahmad S, Abbas O, Hajeer MY, et al. Medical Research Conduct and Publication during Higher Education in Syria: Attitudes, Barriers, Practices, and Possible Solutions. Avicenna J Med. 2022 Jul;12(3):127–37.

17. Bdaiwi Y, Rayes D, Sabouni A, Murad L, Fouad F, Zakaria W, et al. Challenges of providing healthcare worker education and training in protracted conflict: a focus on non-government controlled areas in north west Syria. Confl Health. 2020;14:42.

18. Camargo CP, Tempski PZ, Busnardo FF, Martins M de A, Gemperli R. Online learning and COVID-19: a meta-synthesis analysis. Clin Sao Paulo Braz. 2020;75:e2286.

19. Qazi A, Naseer K, Qazi J, AlSalman H, Naseem U, Yang S, et al. Conventional to online education during COVID-19 pandemic: Do develop and underdeveloped nations cope alike. Child Youth Serv Rev. 2020 Dec;119:105582.

20. NIH Clinical Center News: April 2017 [Internet]. [cited 2023 Jun 3]. Available from: https://clinicalcenter.nih.gov/about/news/newsletter/2017/apr2017/story-april-06-syria.html

21. Hanafi I, Kheder K, Sabouni R, Rahmeh A, Alsalkini M, Hanafi M, et al. Factors Influencing Research Productivity among Syrian Medical Professionals amidst Conflict: a Case-Control Study. 2024.

22. Larsen CM, Terkelsen AS, Carlsen AMF, Kristensen HK. Methods for teaching evidence-based practice: a scoping review. BMC Med Educ. 2019 Jul 11;19(1):259.

23. Aslam F, Shakir M, Qayyum MA. Why Medical Students Are Crucial to the Future of Research in South Asia. PLOS Med. 2005 Nov 29;2(11):e322.

24. Negida AS, Negida A. Egypt’s Premier Medical Student Research Group: A New Model for Medical Student Research in Developing Countries. Cureus [Internet]. 2018 Nov 8 [cited 2023 Jun 3];10(11). Available from: https://www.cureus.com/articles/10927-egypts-premier-medical-student-research-group-a-new-model-for-medical-student-research-in-developing-countries

25. El Achi N, Papamichail A, Rizk A, Lindsay H, Menassa M, Abdul-Khalek RA, et al. A conceptual framework for capacity strengthening of health research in conflict: the case of the Middle East and North Africa region. Glob Health. 2019 Nov 28;15(1):81.

26. Kenjrawi Y, Dashash M. The First Asynchronous Online Evidence-Based Medicine Course for Syrian Health Workforce: Effectiveness and Feasibility Pilot Study. JMIR Form Res [Internet]. 2022 Oct [cited 2023 Jun 3];6(10). Available from: https://www.ncbi.nlm.nih.gov/pmc/articles/PMC9644249/

27. Sabouni A, Chaar A, Bdaiwi Y, Masrani A, Abolaban H, Alahdab F, et al. An online academic writing and publishing skills course: Help Syrians find their voice. Avicenna J Med. 2017 Jul;07(03):103–9.

28. Alabdullah MN, Alabdullah H, Kamel S. Knowledge, attitude, and practice of evidence-based medicine among resident physicians in hospitals of Syria: a cross-sectional study. BMC Med Educ. 2022 Nov 14;22(1):785.

29. Assar A, Matar SG, Hasabo EA, Elsayed SM, Zaazouee MS, Hamdallah A, et al. Knowledge, attitudes, practices and perceived barriers towards research in undergraduate medical students of six Arab countries. BMC Med Educ. 2022 Jan 18;22(1):44.

30. Chun KH, Park WK, Lee SS, Park YS, Kang E. A study on the educational climate, self-directed learning and creative thinking in medical school. Korean J Think Dev. 2010;6(1):179–200.

31. Lee HJ, Kang YJ, Lee SH, Lin Y, Kim DH, Ihm J. Relationship matters: a qualitative study of medical students’ experiences in a learner-driven research program in South Korea. BMC Med Educ. 2023 May 16;23(1):337.

32. Archer A, Berry I, Bajwa U, Kalda R, Di Ruggiero E. Preferred modalities for delivering continuing education to the public health workforce: a scoping review. Health Promot Chronic Dis Prev Can Res Policy Pract. 2020 Apr;40(4):116–25.

33. Mugabo L, Rouleau D, Odhiambo J, Nisingizwe MP, Amoroso C, Barebwanuwe P, et al. Approaches and impact of non-academic research capacity strengthening training models in sub-Saharan Africa: a systematic review. Health Res Policy Syst. 2015 Jun 9;13(1):30.

34. Zhao F, Fu Y, Zhang QJ, Zhou Y, Ge PF, Huang HX, et al. The comparison of teaching efficiency between massive open online courses and traditional courses in medicine education: a systematic review and meta-analysis. Ann Transl Med. 2018 Dec;6(23):458– 458.

35. Launois P, Maher D, Certain E, Ross B, Penkunas MJ. Implementation research training for learners in low- and middle-income countries: evaluating behaviour change after participating in a massive open online course. Health Res Policy Syst. 2021 Apr 6;19(1):59.

36. Li T, Yang N. Comparing MOOCs with Traditional Courses for Quality Teaching in Higher Education. DEStech Trans Soc Sci Educ Hum Sci. 2019 Jan 24;

37. Kop R, Fournier H, Mak JSF. A pedagogy of abundance or a pedagogy to support human beings? Participant support on massive open online courses. Int Rev Res Open Distrib Learn. 2011;12(7):74–93.

38. McMichael C, Waters E, Volmink J. Evidence-based public health: what does it offer developing countries? J Public Health. 2005 Jun 1;27(2):215–21.

39. Fleming-Nouri A, Crocombe D, Sammaraiee Y. Twelve tips on setting up and running a peer-led medical education society. Med Teach. 2016 Dec 1;38(12):1199–203.

40. Allikmets S, Vink JP. The benefits of peer-led teaching in medical education. Adv Med Educ Pract. 2016 May 31;7:329–30.

41. Ahmadi SF, Baradaran HR, Ahmadi E. Effectiveness of teaching evidence-based medicine to undergraduate medical students: a BEME systematic review. Med Teach. 2015 Jan;37(1):21–30.

